# Reliability and Structure of Diabetes Diet Adherence Scale (D-DAS): A Follow-up Study among Type 2 Diabetes Patients of India

**DOI:** 10.1101/2024.05.25.24307586

**Authors:** Savitesh Kushwaha, Rachana Srivastava, Sanjay Kumar Bhadada, Jarnail Singh Thakur, Vivek Sagar, Rupa Tharu, Poonam Khanna

**Affiliations:** Department of Community Medicine & School of Public Health, PGIMER, Chandigarh, India; Department of Endocrinology, PGIMER, Chandigarh, India

**Keywords:** Adherence, Diabetes, Diet, Scale, Validation

## Abstract

**Introduction:** Measuring dietary adherence is essential while prescribing a diet plan for type 2 diabetes. This study aims to develop and validate a diabetes diet adherence scale (D-DAS) among type 2 diabetes patients in India.

**Methods:** A four-month non-randomised follow-up study was conducted among (n=120) type 2 diabetes patients attending the outpatient clinic. The adherence to the prescribed diet plan was determined using the D-DAS scale. Multiple correspondence analysis (MCA) was used to determine the underlying dimensions of the scale, and confirmatory factor analysis (CFA) with multiple reliability measures was used to determine the reliability and construct validity of the scale. The latent class analysis (LCA) was done to determine the cutoffs for the scale. Although the sample size underpowered for CFA and LCA stability.

**Results:** The CFA indicated high reliability, with McDonald’s ω = 0.836 and Guttman’s λ2 = 0.836. The MCA identified dimensions that accounted for 68.4% of the total variance. The D-DAS scale further demonstrated a robust latent structure, evidenced by a composite reliability of 0.900 and an average variance extracted (AVE) of 0.565. Regarding adherence outcomes, 68.3% of participants exhibited adherence to the prescribed diet plan, whereas 31.7% demonstrated non-adherence.

**Conclusion:** D-DAS is a reliable and valid scale for measuring dietary adherence among type 2 diabetes patients in India. Adherence was moderate, suggesting the need for targeted interventions to improve dietary compliance and diabetes management.

## Introduction

Worldwide, a considerable amount of the healthcare budget goes towards paying for diabetes treatment. The quality and amount of diet impact the long-term prevention and control of diabetes and its consequences.(1) Nutrition therapy has always been a recognized cornerstone in the management of diabetes. Adopting dietary recommendations for diabetes is essential for patients with diabetes to maintain glucose control and achieve long-term health goals (2). Dietary changes can reduce glycated haemoglobin (HbA1c) levels by 1% to 2%, making them one of the key treatment components (3). WHO advises patients to maintain a healthy body weight, engage in regular physical activity for at least 30 minutes and moderate-intensity activity on most days, follow a healthy diet, abstain from tobacco products, and achieve and maintain a healthy weight to prevent type 2 diabetes and its complications. A good diet helps prevent non-communicable diseases (NCDs), such as diabetes, heart disease, stroke, and cancer, as well as malnutrition in all manifestations (4). Following a diabetic diet has also been demonstrated to reduce blood pressure, normalize lipid irregularities, and enhance glucose levels, all connected to the micro- and macro-vascular issues of diabetes (5).

Although dietary modification has been anticipated as the keystone of type 2 diabetes management and is typically recommended as the first step, it is considered one of the most challenging aspects of diabetes management. Regularly implementing recommended dietary practices for individuals with type 2 diabetes requires collaboration between the patient and the healthcare provider. Despite the formulation of comprehensive guidelines for achieving optimal diabetes care, evidence indicates that most patients with diabetes have difficulty incorporating nutrition recommendations into their everyday lives (6). Factors identified for poor adherence to dietary recommendations include socioeconomic status, duration of diabetes, and diabetes severity (4). However, switching to a healthy diet requires giving up decades-long eating habits and the numerous little behaviours associated with them throughout each day. The amorphous and intricate influences of the psychological, emotional, and societal difficulties related to diet adherence may be felt throughout the day (7).

The ICMR recommends six small meals daily for all patients with diabetes, calorie distribution, up to 55–60% of energy from complex carbs, and proteins should make up 12–15% of total energy consumption of a patients with diabetes. Consuming proteins from lean meats, fish, low-fat dairy products, pulses, soy, grains, and peas is advised. 20 to 30 percent of daily energy intake should come from fats (8). While providing clear recommendations on healthy eating patterns for type 2 diabetes management, the ADA approach recognizes the complexities of dietary adherence and encourages healthcare providers to focus on individualized care and addressing the barriers individuals face in adopting and maintaining these dietary changes (9). Earlier studies also indicate that many patients do not follow their recommended diets. Although the extent of T2D diet non-compliance is unknown, research indicates that this extent ranges from 2.2% to 87.5% (10). Developing interventional programmes to encourage healthy eating might greatly benefit from understanding the dimensions of adherence to a healthy diet in diabetes patients, particularly from a healthcare professionals (HCPs) viewpoint.

The scale to measure adherence to a prescribed diet plan in diabetes was not available for the Indian population. Therefore, the present study aims to translate and validate the Diabetes Diet Adherence Scale (11) among the Indian population and determine the underlying dimensions of the scale contributing towards adherence and non-adherence.

## Methods

This was a four-month, single-group, non-randomized follow-up study conducted among patients with type 2 diabetes mellitus attending the outpatient clinic of the Endocrine Department, PGIMER, Chandigarh, following the TRENDS statement checklist (12). The study used random sampling to select participants at baseline. We have determined the sample size using G-power software based on a previous study (13) among 218 Indian T2DM patients (effect size=0.564). The estimated sample size was 102. The final calculated sample size with 80% power, including 10% follow-up loss, was 120. One hundred twenty patients were recruited at the baseline, and at the endline, data from 104 patients were collected (14% attrition). Demographic information was collected encompassing age, gender, smoking status, alcohol consumption, food habits, marital status, work status, and educational attainment. Body weight was measured using a digital weighing scale (Dr Trust), while height assessment was conducted with a height board (SECA), adhering to standardized procedures. Fasting blood glucose (FBG) values were obtained at both baseline and endline utilizing a glucometer (Dr Trust) according to the standard protocol established by the Department of Endocrinology, PGIMER, Chandigarh. Glycated haemoglobin (HbA1c) levels were extracted from the patients’ outpatient department (OPD) cards and laboratory reports, ensuring that results were not older than one month at the time of data collection.

### Diabetes Diet Adherence Scale (D-DAS) and Intervention

The original (English) and translated (Hindi) diabetes diet adherence scale (D-DAS) (Mohammed et al., 2019) (11) was used to collect data on adherence. The permission to use these scales was obtained from the corresponding author of Mohammed et al., 2019. The D-DAS was translated into Hindi following internationally recommended guidelines for cultural adaptation of psychometric instruments (14). The translation process involved (i) independent forward translation by two bilingual experts, (ii) reconciliation and synthesis by a multidisciplinary expert panel for conceptual and cultural appropriateness, (iii) independent back-translation, (iv) review of the back-translated version for equivalence to the original, and (v) pilot testing (n=10) with cognitive debriefing in a sample of the target population to ensure clarity, relevance, and conceptual equivalence of all items. Feedback informed final refinements to maximize both content validity and cross-cultural applicability. The D-DAS scale was developed in accordance with Morisky 8 item Medication Adherence Scale and consists of 10 questions about prescribed dietary adherence in diabetes (Suppl_File_Table_1). The diet plan prescribed to the patients was based on the recommendations for diabetes from the Indian Council of Medical Research (8). The diet plan was made considering the accessibility and affordability of the food products and the minimum extra burden on pocket expenditure by expert dietitians. The patients diligently adhered to the diet plan, ensuring compliance throughout the study. Monthly phone calls were made to each patient to monitor their progress and provide support. These phone calls served as a means of check-ins, allowing the medical team to address any concerns or challenges faced by the patients and provide guidance and motivation to stay on track. In addition to the phone calls, the patients also received regular text messages twice a month. These messages were carefully crafted to provide reminders, encouragement, and helpful tips related to their dietary requirements. The D-DAS scale was employed after the completion of 4-month follow-up period.

### Statistical Analysis

The Mardia’s test for multivariate normality was conducted to determine the normal distribution of the data. The descriptive analysis was used to report the percentages and Median with IQR. The MCA (Multiple Correspondence Analysis) was performed using the ‘indicator’ method to identify the underlying dimensions that structure the relationships between the several categorical variables. The squared cosine (cos2) assesses the quality of representation of the categories or levels of a categorical variable on the dimensions of the analysis. The eta-squared (ETA^2^) was based on the analysis of variance (ANOVA) framework and represents the proportion of variance in the observed variables explained by the latent construct, similar to the explained variance in regression analysis. The psychometric properties of the adherence scale were further evaluated using Two-Parameter Logistic (2PL) Item Response Theory (IRT) analysis to assess item discrimination and difficulty parameters. We have conducted Confirmatory Factor Analysis (CFA) using a weighted least squares mean, and variance adjusted (WLSMV) estimator, which is useful in construct validation of categorical variables because it allows researchers to estimate the degree to which individual items are influenced by the general factor versus specific factors. Our data don’t represent equal factor loadings (tau equivalence) and normality, so the reliability measurement using Cronbach’s α coefficient might not be reliable. Cronbach’s α assumes that all scale items contribute equally to the underlying construct (tau-equivalence) and that errors are uncorrelated, which is often not the case in practical psychometric applications. The obtained p-value was <0.001, indicating a highly significant difference in model fit between the congeneric and tau-equivalent models. The assumption for all items measures the latent construct with equal loadings (tau-equivalence) was violated in our data. These restrictive assumptions can lead to either underestimation or overestimation of the true internal consistency (15). In contrast, McDonald’s ω and Guttman’s λ2 account for differences in item loadings and allow for more realistic estimation of the reliability in scales where items vary in their strength of association with the latent construct (16). Therefore, we have also used McDonald’s Omega (ω) and Gutmann’s Lambda-2 to determine the reliability and internal consistency. The variables selected for analysis in Cronbach’s α, McDonald Omega (ω) and Gutmann’s Lambda-2 were selected based on factor loading value (>0.50) in CFA. The varimax rotation was used to maximise and simplify the loadings of factors. The AVE (Average Variance Explained) Composite Reliability (CR) values were calculated according to the equations given by Fornell and Larcker (17):

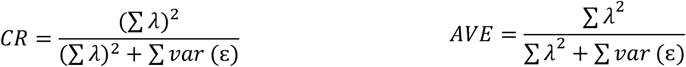

Where, λ = factor loading, ε = (1-λ^2^)

Following construct validation of the D-DAS and selection of 7 items based on factor loadings, eta^2^ values, and reliability parameters, Latent Class Analysis (LCA) was performed on the validated 7-item dietary adherence scale to identify distinct adherence profiles. The optimal number of classes was determined by comparing models with different numbers of classes using the Bayesian Information Criterion (BIC), with a 2-class solution providing the best fit. Classes were labelled as high and low adherence based on obtained cutoff values from their mean dietary adherence scores and item response probabilities. Cutoff values were then derived from the boundaries between these LCA-identified groups. Statistical significance was set at p < 0.05 for assumption testing (normality) and primary outcome analyses. For exploratory group comparisons examining adherence rates across demographic subgroups, Bonferroni correction for multiple testing was applied, resulting in an adjusted significance threshold of p < 0.0125. All the analysis was conducted in R version 4.0.2 with packages (*FactoMineR, factoextra, lavaan, semTools, ggplot2, psych, poLCA, mirt etc*.) and JASP software version 0.15. The informed and written consent were obtained from the participants. Standardized interviewer training, confidentiality assurance and self-administering questionnaire were used to minimize the interviewer bias. The ethical clearance for this study was obtained from the Institutional Ethics Committee, PGIMER, Chandigarh.

## Results

The distribution of demographic variables includes 64.17% of males and 35.83% of females. 2.50% were smokers, and 23.23% consumed alcohol. The vegetarian (60.00%) group was highest compared to non-vegetarian (39.17%) and ovo-vegetarian (0.83%) groups (Suppl_File_Table_2). The Mardia’s test suggests that in the skewness variable, the beta-hat (estimated population parameter) was 0.589, the kappa value was 10.214, and the p-value was 0.037. This suggests a non-normal distribution, as the p-value was below the typical significance threshold of 0.05. For the kurtosis variable, the beta-hat was 6.53, the kappa value was -1.874, and the p-value was 0.061. This result was less clear, as the p-value was above the 0.05 threshold but still relatively low. The positive value of beta-hat suggests that the distribution of this variable peaks more than a normal distribution. In contrast, the negative kappa value suggests that there may be some non-normality present. Overall, Mardia’s test suggests that there may be some non-normality in the distribution of both variables, particularly for the skewness variable (Suppl_File_Table_3). The median age, FBG, HbA1c, and BMI value at the baseline was 59.90 years (IQR: 52.00-62.25), 130.00 mg/dL (IQR: 112.00-176.50), 8.40% (IQR:8.30-9.00) and 25.16 (IQR: 22.23-28.18), respectively (Suppl_File_Table_4).

The Multiple Correspondence Analysis was conducted to identify the underlying dimensions that structure the relationships between the several categorical variables. The scree plot shows that dimension-1 to dimension-4 account for 68.4% of the total variance in the adherence scale (Figure 1). Based on the eigenvalues obtained in the scree-plot, we have retained four dimensions with explained variance between the dependent (questions) and independent variable (diet plan). The dimension-1 has the highest eigenvalue, and the squared eta values were >0.20 for D1 to D7 (directly related to the prescribed diet plan). Dimension 1 primarily separates endorsements of non-adherence behaviours (‘Yes’ on D1–D7) on the positive side from endorsements of adherence (‘No’ on D1–D7) on the negative side, consistent with item coding (No = adherence; Yes = non-adherence), and is therefore labelled non-adherence behaviours vs adherence to the prescribed plan (Suppl_File_Fig_1). Categories from D1–D7 demonstrate substantial contributions to Dimension 1, with D2 showing the highest contribution (11.3%) and exceptional representation quality (cos^2^ = 0.661), indicating that implementation-lapse items collectively define this primary axis. Variables D1 (cos^2^ = 0.260), D3 (cos^2^ = 0.443), D4 (cos^2^ = 0.511), D5 (cos^2^ = 0.373), D6 (cos^2^ = 0.257), and D7 (cos^2^ = 0.477) also display acceptable-to-good representation quality on Dimension 1. The variable association measures (η^2^) further confirm this pattern, with D2 demonstrating the strongest association with Dimension 1 (η^2^ = 0.661), followed by D4 (η^2^ = 0.511) and D7 (η^2^ = 0.477). This dimensional structure aligns closely with the CFA framework, where D1–D7 exhibited moderate-to-strong factor loadings, thereby supporting construct coherence between the MCA-derived structure and the validated D-DAS implementation-lapse factor. The convergent evidence from both contribution percentages and representation quality metrics reinforces the theoretical integrity of this dietary adherence dimension (Figure 2).

**Figure 1.**
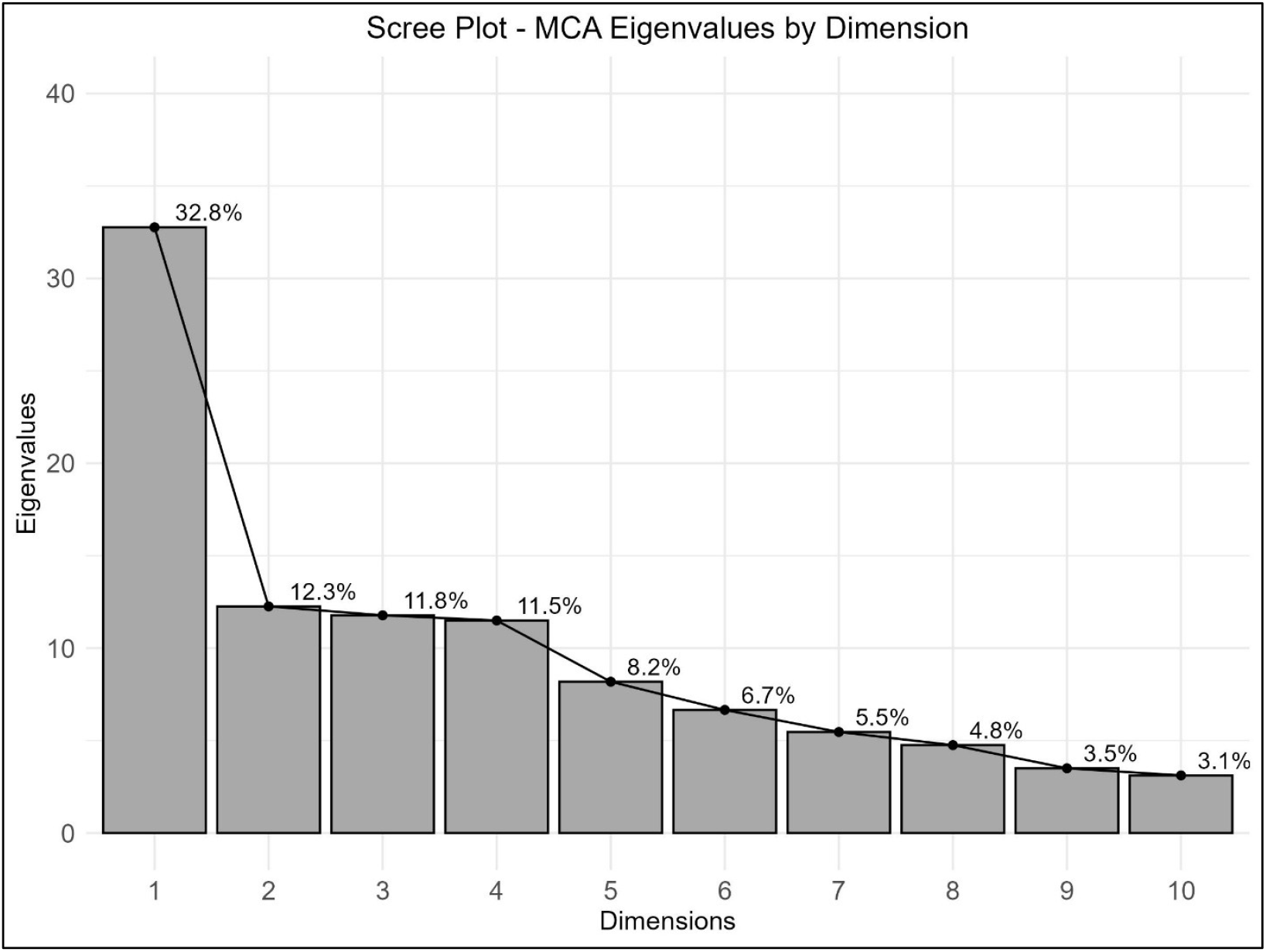
Scree plot showing the eigen values with percentage of explained variance in various identified dimensions.

**Figure 2.**
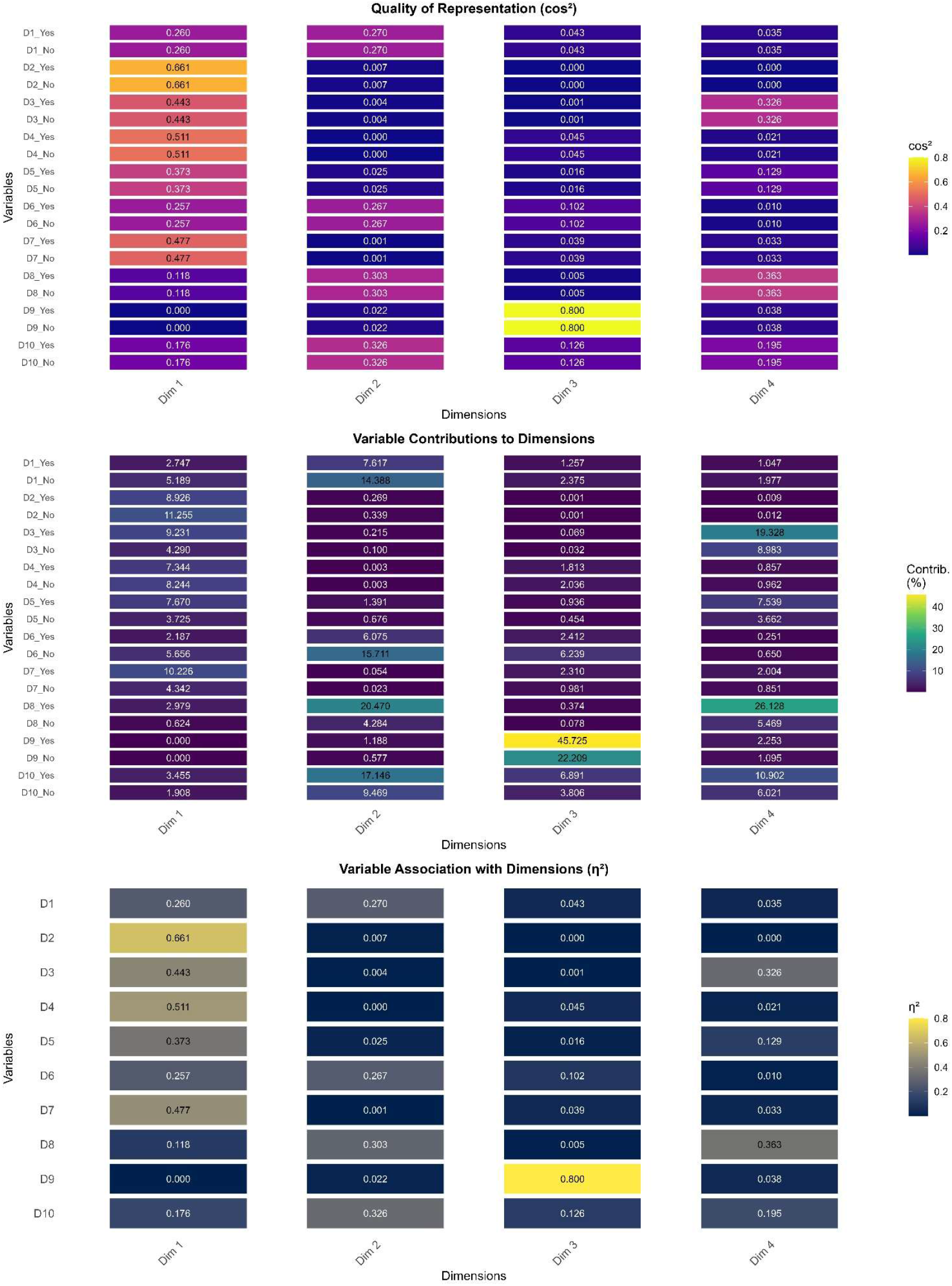
Heatmap showing cos2 distribution, contribution and squared eta for dimension 1 to 4.

The variables D7, D3, D5, D4 and D2 were associated with each other and moderately to strongly associated with dimension 1, whereas the variable D9 was strongly associated with dimension 3. The association between a variable and a dimension reflects how much of the variability can be explained by the corresponding dimensions. The plot for two-dimension (all six pairs) combinations was provided in Suppl_File_Fig_2.

Standardized residual covariance diagnostics showed predominantly small residuals, with a small number of item pairs exhibiting substantial local misfit (|residual| > 2.58)(18), most representing D1–D10, D3–D8, and D6–D10. These localized discrepancies suggest potential unmodelled item dependencies and are reported as diagnostic findings rather than addressed through post-hoc model modification (Suppl_File_Fig_3).

The 2PL IRT analysis provided item-level psychometric assessment, with critical differences observed between item sets. The 7-item model (D1-D7) achieved successful convergence after 42 iterations (log-likelihood = -389.14), demonstrating stable parameter estimation. Discrimination parameters ranged from 1.29 to 5.61, with four items (D2, D3, D4, D7) showing excellent discrimination (ai > 2.0) and three items (D1, D5, D6) displaying acceptable discrimination (a_1_ > 1.2). Difficulty parameters indicated appropriate coverage across the adherence continuum, from easily endorsed behaviours (D2, D6) to more challenging adherence practices (D3, D7).

Conversely, the full 10-item model failed to converge after 500 iterations, with items D8, D9, and D10 exhibiting poor discrimination properties (ai < 1.0). The convergent findings from both MCA dimensional analysis and IRT psychometric evaluation support the retention of a refined 7-item scale, which demonstrates superior measurement stability and validity for assessing dietary adherence behaviours (Suppl_File_Fig_4).

### Reliability and Construct Validity

The one-factor solution was developed using the confirmatory factor analysis. In the CFA, question D2 has a high loading of 0.962, indicating that it is strongly related to the factor. Questions D1, D3, D4, D5, D6, and D7 also have moderate to strong loadings, indicating that they are also related to the factor. Questions D8, D9, and D10 have lower loadings, indicating weaker relationships with the factor (Table 1). The confirmatory factor analysis demonstrated good overall model fit for the single-factor structure. The Comparative Fit Index (CFI) (0.981) and Tucker-Lewis Index (TLI) (0.975) both exceeded the recommended threshold of 0.95, indicating that the proposed model fits substantially better than a baseline independence model. The Root Mean Square Error of Approximation (RMSEA) value of 0.056 falls within the acceptable range (< 0.08), suggesting reasonable approximation error in the population. However, the Standardized Root Mean Square Residual (SRMR) value of 0.144 is concerning as it exceeds the recommended cutoff of 0.08 (19, 20), indicating that the model may not adequately reproduce the observed correlations among items (Table 2). This suggests there may be some local misfit in the model - perhaps some items share residual variance not captured by the single factor, or there may be method effects or secondary dimensions present. However, even after using 2-factor model (Factor-1=D1-D7; Factor-2=D8-D10) no major change in SRMR (0.137) was observed. The AVE coefficient of 0.565 suggests that the factor explains about 56% of the variance in the manifest variables (D1 to D7). The CR coefficient of 0.900 indicates that the factor is highly reliable and that the manifest variables (D1 to D7) are highly inter-correlated. The high AVE and CR values suggest that the factor adequately represents the underlying construct and that the manifest variables are reliable indicators of the construct (Table 2).

**Table 1.**
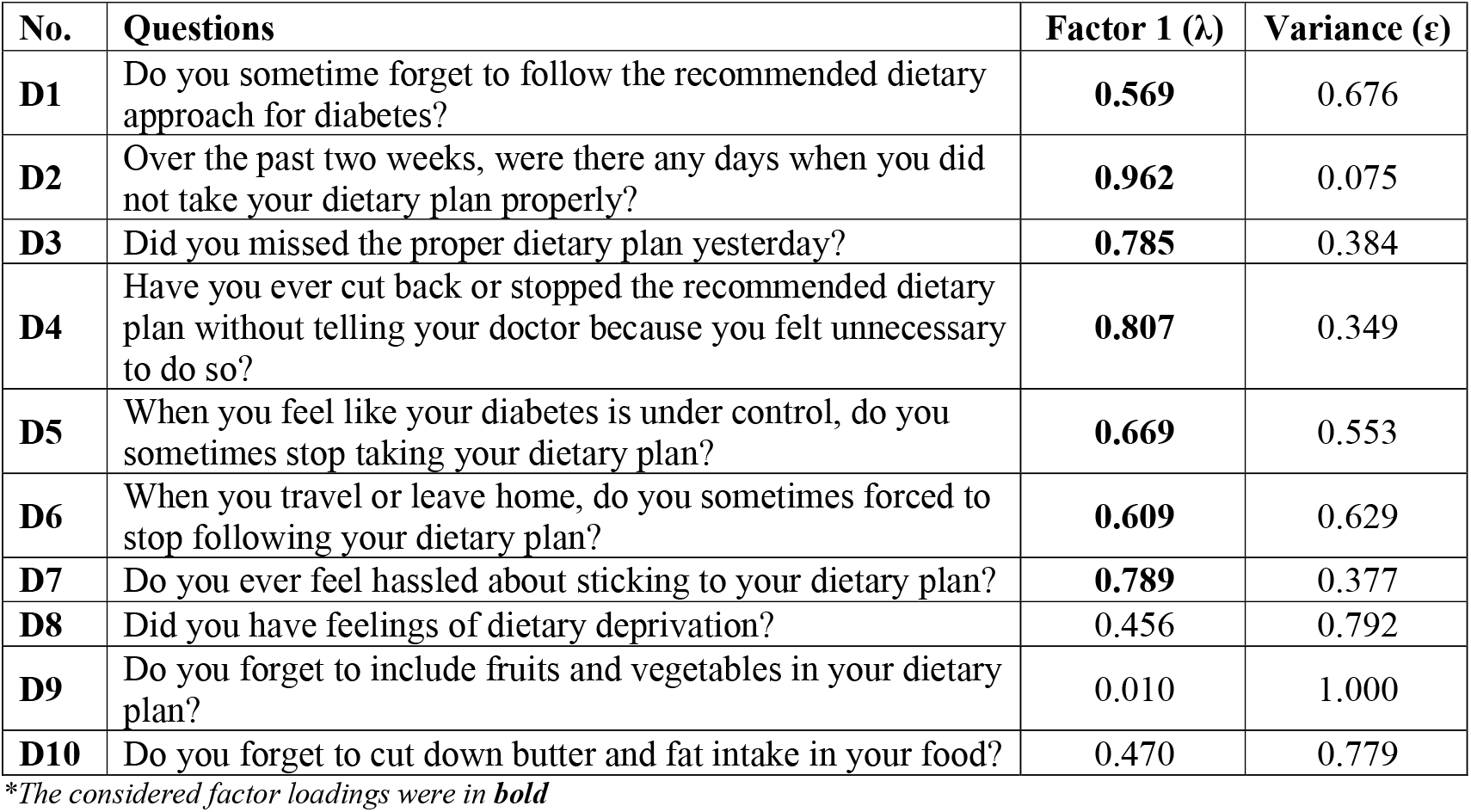
The factor loadings obtained for factor-1 in CFA.

| No. | Questions | Factor 1 ($\lambda$) | Variance ($\epsilon$) |
| --- | --- | --- | --- |
| **D1** | Do you sometime forget to follow the recommended dietary approach for diabetes? | **0.569** | 0.676 |
| **D2** | Over the past two weeks, were there any days when you did not take your dietary plan properly? | **0.962** | 0.075 |
| **D3** | Did you missed the proper dietary plan yesterday? | **0.785** | 0.384 |
| **D4** | Have you ever cut back or stopped the recommended dietary plan without telling your doctor because you felt unnecessary to do so? | **0.807** | 0.349 |
| **D5** | When you feel like your diabetes is under control, do you sometimes stop taking your dietary plan? | **0.669** | 0.553 |
| **D6** | When you travel or leave home, do you sometimes forced to stop following your dietary plan? | **0.609** | 0.629 |
| **D7** | Do you ever feel hassled about sticking to your dietary plan? | **0.789** | 0.377 |
| **D8** | Did you have feelings of dietary deprivation? | 0.456 | 0.792 |
| **D9** | Do you forget to include fruits and vegetables in your dietary plan? | 0.010 | 1.000 |
| **D10** | Do you forget to cut down butter and fat intake in your food? | 0.470 | 0.779 |

\*The considered factor loadings were in **bold**

**Table 2.** The CFI, TLI, RMSEA, SRMR showing model fit estimates and AVE and CR measures for construct validation of the scale.

| Measures | Values |
| --- | --- |
| Comparative Fit Index (CFI) | 0.981 |
| Tucker-Lewis Index (TLI) | 0.975 |
| Root Mean Square Error of Approximation (RMSEA) | 0.056 |
| Standardized Root Mean Square Residual (SRMR) | 0.144 |
| Average Variance Extracted (AVE) | 0.565 |
| Composite Reliability (CR) | 0.900 |

The reliability analysis results indicate that the scale has high internal consistency reliability. McDonald’s omega (ω) was 0.836, indicating that 83.6% of the variance in observed scores was attributable to true score variance after accounting for measurement error. Cronbach’s alpha (α) was also high at 0.817, though McDonald’s omega is considered more appropriate given that tau-equivalence assumptions were not met in our dataset. The measures for Factor-1 show higher reliability and internal consistency than individual scale items (Table 3).

**Table 3.**
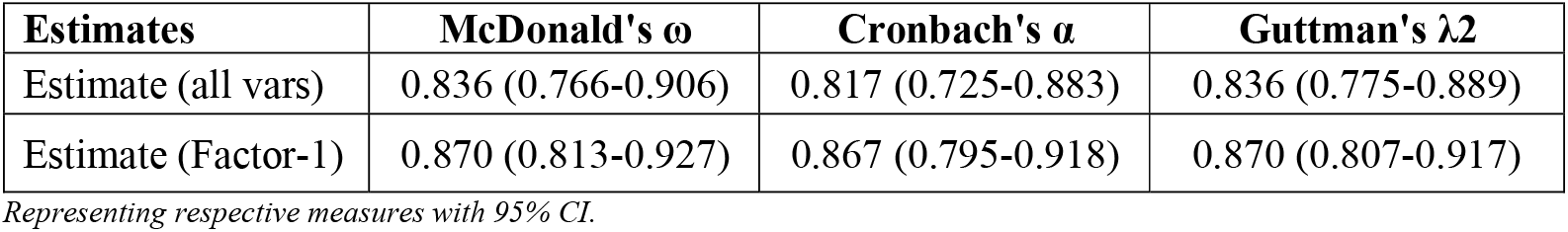
The assessment of reliability and internal consistency of the diet adherence scale.

The description of LCA summary with Item Response Probabilities of D1 to D7 were provided in supplementary file table 5. The adherence results show 68.3% of participants shown high adherence to the prescribed diet plan (Table 4). The adherence across gender, alcohol consumption, smoking and BMI was not significant (Suppl_File_Table_6).

**Table 4.** The overall mean, median and percent of adherence and non-adherence to the prescribed diet *(Cutoff: 3*.*00)*.

| Measures | Score (Mean±SD) | Score (Median IQR) | n | Percent (%) |
| --- | --- | --- | --- | --- |
| High Adherence | 4.83±1.39 | 5.0 (4-6) | 71 | 68.3 |
| Low Adherence | 0.94±0.79 | 1.0 (0-2) | 33 | 31.7 |

## Discussion

Our study successfully validated the D-DAS scale for estimating the adherence to the prescribed diet plan among the patients with diabetes in India. The fundamental set of diabetes care includes self-management education and support (DSMES), medical nutrition therapy (MNT), physical activity, smoking cessation counselling, and psychological care (21). The American Diabetes Association emphasises that MNT is the base of the diabetes management plan, and the requirement for MNT should be re-evaluated regularly by healthcare personnel. Collaborating with people with diabetes and paying special attention to their changing health status and life stages can be a significant approach to managing type 2 diabetes (22). In India, there is a propensity to consume calorie-dense foods at the expense of a variety of foods, which may lead to micronutrient shortages as well as the emergence of type 2 diabetes and other related metabolic illnesses (23). An umbrella meta-analysis mentions no evidence to favour any specific macronutrient composition or dietary style over others in published meta-analyses of hypocaloric diets for weight management in adults with type 2 diabetes. The most successful methods seem to be very low-calorie diets and formula meal replacement, which often provide less energy than self-administered food-based diets (24). Another meta-analysis suggests that whole grains positively impact glucose metabolism (25). However, the food processing and preparation method was important to maintain the structural integrity of the whole grains (26, 27). Working groups from India, including the Indian Council of Medical Research (ICMR) and the Research Society for the Study of Diabetes in India (RSSDI), have framed treatment recommendations for diabetes using nutritional principles. The RSSDI supports comprehensive lifestyle changes, including MNT, to help patients achieve their ideal glycemic and lipemic indexes and improve their general well-being (28, 29).

The impact of MNT on diabetes strongly depends on adherence and compliance with the MNT. There were several measurement scales available for different management components. The ADQ (Adherence in Diabetes Questionnaire) scale determines adherence to insulin injections (30). Another scale determines adherence to the Mediterranean diet (31, 32), and Diet Quality Index-International (DQI-I) is applicable for diet quality comparisons across countries (33). The Perceived Dietary Adherence Questionnaire (PDAQ) was developed based on a Canadian food guide (34) and was not validated in India (35). The scales for adherence to lifestyle advice (36) and multiple scales for measuring medication adherence were available (37-39). For physical activity, the Global Physical Activity Questionnaire (GPAQ) (40), International Physical Activity Questionnaire (IPAQ) (41), International Physical Activity Questionnaire-Short Form (IPAQ-SF) (42) and Exercise Adherence Rating Scale (EARS) (43) were used in different research fields.

However, it has been observed that MNT adherence was medium to poor in most studies (44-46). The scales for measuring dietary adherence among diabetes patients were available (47, 48), but no validated scale was available to measure the prescribed dietary adherence among diabetes patients in India. In the present study, the underlying dimensions and structure of the D-DAS scale suggest that dimension-1 consists of the questions from D1 to D7, which have a good quality of representation and strong correlation, suggesting similarity in the structure of the scale. The confirmatory factor analysis also shows that the underlying construct follows the MCA dimensions, and factor-1, which includes D1 to D7, shows strong factor loadings. The measure of construct validity suggests that the D-DAS scale confers a valid underlying construct and can be used for measuring adherence to diabetes diet plan/MNT in the Indian population. The D8 to D10 was not directly related to the prescribed diet plan, as it was related to fruit, vegetable and fat intakes. Rephrasing D8–D10 to anchor behaviours explicitly to the prescribed MNT plan and verifiable, time-bound adherence acts (e.g., ‘On days you follow the prescribed plan, do you meet the fruit/vegetable targets specified in your plan?’), thereby improving construct match and expected loading. A recently published study using the same scale in Ghana showed 70.6% adherence to the dietary regime (49). Adherence to dietary restrictions, for instance, depends on the market’s ongoing supply of affordable dietary alternatives, their accessibility and affordability to the patient, and the patient’s motivation for adherence and the provision of appropriate dietary advice. The low dietary adherence might also be due to a lack of accessibility, cultural inappropriateness, or difficulty in preparation. The Knowledge, Attitude and Practice-based intervention models can help increase dietary adherence among diabetes patients (50, 51). To ensure reduced costs and more access to good foods, as well as the contrary for those posing an increased risk to health, these will necessitate readjusting national or state policies for food procurement, pricing, and marketing.

Integration of the D-DAS into routine clinical practice, holds significant promise for improving diabetes care at both individual and systemic levels. By providing a reliable and valid tool to assess diet adherence, the D-DAS facilitates the early identification of individuals with dietary non-adherence. Utilization of this scale within routine clinical encounters has the potential to foster patient engagement, promote shared decision-making, and support the setting of realistic, personalized dietary goals. In the initial consultation phase after prescribing a diet plan, healthcare providers can identify adherence challenges. The high implementation-lapse scores (questions 1-7) may indicate practical barriers requiring problem-solving interventions, while elevated behavioral-inconsistency scores (questions 8-10) might suggest motivational or self-regulatory concerns warranting different therapeutic approaches. The structured assessment provided by D-DAS can also enhance follow-up consultations by allowing both clinicians and patients to monitor changes in adherence behavior systematically. The increasing integration of digital health tools offers opportunities to facilitate the implementation of the D-DAS scale in busy outpatient settings. The use of QR codes, tablet-based interfaces, and automated processing of scanned questionnaires using large language models (LLMs) can enable rapid and efficient assessment of participants. Furthermore, as the D-DAS is designed to be self-administered, it minimizes the need for additional personnel support during data collection. To enhance accessibility among illiterate or low-literacy participants, tablet-based platforms can be equipped with voice-over features to assist respondents in understanding and selecting appropriate responses.

With respect to limitations, the use of self-reported measures for dietary adherence, while practical for large surveys, may introduce bias arising from recall inaccuracies or the tendency of participants to provide socially desirable responses. Additionally, the underpowered sample size may limit the ability to establish causal relationships between diet adherence and clinical outcomes such as glycemic control. Future studies should aim to use large and multi-center samples to enhance both the interpretability and validity (CFA and LCA) of adherence classification of the D-DAS.

## Conclusion

The findings of this study underscore that adherence to the prescribed medical nutrition therapy (MNT) among patients with type 2 diabetes was generally suboptimal. Such limited adherence poses challenges in achieving adequate control of fasting blood glucose levels through diet alone. The development and validation of the D-DAS scale confirmed a reliable and valid construct for assessing dietary adherence to MNT within the Indian population. The results highlight the importance of promoting comprehensive lifestyle modifications, with particular emphasis on MNT, to support patients in attaining optimal glycemic and lipidemic profiles and enhancing overall well-being. Future research should prioritize the design and evaluation of targeted interventions to improve adherence to MNT, while policymakers may consider implementing measures that strengthen food security for individuals living with diabetes.

## Supporting information

https://drive.google.com/file/d/1SwZWihGjBh-dR-WjsNAQCmhvH-BcSS0G/view?usp=sharing

## Data Availability

All data produced in the present study are available upon reasonable request to the authors

## Acknowledgements

We would like to thank the participants involved in this study and the health workers who assisted with the data collection. We would like to thank the authors (Mohammed MA et al., 2019) of D-DAS scale for provide us permission to use the scale.

## Funding and Assistance

This manuscript is not funded by any organisations or institutions.

## Author Contributions Statement

PK and SK designed the manuscript. SK and RT did the data cleaning and sorting of variables. SK and RS performed all statistical analyses. PK, JST, and SKB interpreted the results. RT and SK developed all the figures. PK and VS wrote the manuscript. SKB, JST and PK reviewed the manuscript. All the authors had the decision to submit it for publication.

## Data Sharing

Data described in the article will be made available upon reasonable request from the corresponding author. The D-DAS scale (English and Hindi Version) can be used after sending an email to any authors.

