## Supplementary material for "Reliability and Structure of Diabetes Diet Adherence Scale (D-DAS): A Follow-up Study among Type 2 Diabetes Patients of India": https://drive.google.com/file/d/1SwZWihGjBh-dR-WjsNAQCmhvH-BcSS0G/view?usp=sharing

**Supplementary Table 1: The Diabetes Diet Adherence Scale (D-DAS) (English and Hindi).**

| Sl. No. | Questions | Response |
|---|---|---|
| D1. | Do you sometime forget to follow the recommended dietary approach for diabetes? (क्या आप कभी-कभी मधुमेह के लिए अनुशंसित आहार संबंधी दृष्टिकोण का पालन करना भूल जाते हैं?) | Yes/No |
| D2. | Over the past two weeks, were there any days when you did not take your dietary plan properly? (पिछले दो सप्ताहों में क्या कोई ऐसा दिन था जब आपने अपनी आहार योजना का पालन ठीक से नहीं किया?) | Yes/No |
| D3. | Did you missed the proper dietary plan yesterday? (क्या आपने कल उचित आहार योजना का पालन नहीं किया?) | Yes/No |
| D4. | Have you ever cut back or stopped the recommended dietary plan without telling your doctor because you felt unnecessary to do so? (क्या आपने कभी अपने डॉक्टर को बताए बिना अनुशंसित आहार योजना में कटौती की है या उसे बंद कर दिया है, क्योंकि आपको ऐसा करना अनावश्यक लगा?) | Yes/No |
| D5. | When you feel like your diabetes is under control, do you sometimes stop taking your dietary plan? (जब आपको लगता है कि आपका मधुमेह नियंत्रण में है, तो क्या आप कभी-कभी अपनी आहार योजना लेना बंद कर देते हैं?) | Yes/No |
| D6. | When you travel or leave home, do you sometimes forced to stop following your dietary plan? (जब आप यात्रा करते हैं या घर से बाहर जाते हैं, तो क्या कभी-कभी आपको अपनी आहार योजना का पालन करना बंद करना पड़ता है?) | Yes/No |
| D7. | Do you ever feel hassled about sticking to your dietary plan? (क्या आपको कभी अपने आहार योजना पर टिके रहने में परेशानी महसूस होती है?) | Yes/No |
| D8. | Did you have feelings of dietary deprivation? (क्या आपको आहार से वंचित होने का एहसास हुआ?) | Yes/No |
| D9. | Do you forget to include fruits and vegetables in your dietary plan? (क्या आप अपने आहार में फल और सब्जियां शामिल करना भूल जाते हैं?) | Yes/No |
| D10. | Do you forget to cut down butter and fat intake in your food? (क्या आप अपने भोजन में मक्खन और वसा का सेवन कम करना भूल जाते हैं?) | Yes/No |

**Scores: Yes=0; No=1 (Total = 10)**

**Supplementary Table 2: Socio-demographic information of diabetic patients.**

| Variables | Category | N |
|---|---|---|
| Gender | Male | 77 (64.17) |
| | Female | 43 (35.83) |
| Smoking | No | 117 (97.50) |
| | Yes | 3 (2.50) |
| Alcohol | No | 92 (76.76) |
| | Yes | 28 (23.23) |
| Food Habit | Veg | 72 (60.00) |
| | Non-Veg | 47 (39.17) |
| | Ovo | 1 (0.83) |
| Marital Status | Married | 1 (0.83) |
| | Unmarried | 119 (99.17) |
| Work Status | Government Employee | 16 (13.33) |
| | Non-Government Employee | 6 (5.00) |
| | Self Employed | 27 (22.50) |
| | Non-Paid | 2 (1.67) |
| | Student | 30 (25.00) |
| | Homemaker | 36 (30.00) |
| | Retired | 0 (0.00) |
| | Unemployed (able to work) | 3 (2.50) |
| | Unemployed (unable to work) | 0 (0.00) |
| Education | No Formal schooling | 24 (20.00) |
| | Less than primary school | 9 (7.50) |
| | Primary school completed (I to V) | 5 (4.17) |
| | Middle school completed (VI to VIII) | 5 (4.17) |
| | High school completed (IX & X) | 32 (26.67) |
| | Secondary School completed (XI & XII) | 13 (10.83) |
| | Graduate | 27 (22.50) |
| | Postgraduate | 5 (4.17) |

**Supplementary Table 3: Mardia's test for multivariate normality showing non-significant p-values.**

| Measures | Beta-hat | kappa | p-value |
|---|---|---|---|
| Skewness | 0.589 | 10.214 | 0.037 |
| Kurtosis | 6.530 | -1.874 | 0.061 |

**Supplementary Table 4: Showing the median and IQR values of various continuous variables at baseline.**

| Variables | N | Median | IQR |
|---|---|---|---|
| Age (years) | 120 | 59.50 | 52.00 – 62.25 |
| FBS (mg/dL) | 120 | 130.00 | 112.00 – 176.50 |
| HbA1c (%) | 66* | 8.40 | 8.30 – 9.00 |
| Weight (Kg) | 120 | 65.00 | 60.00 – 73.25 |
| Height (cm) | 120 | 162.80 | 156.00 – 170.70 |
| BMI | 120 | 25.16 | 22.23 – 28.18 |

**Record based measurement*

**Supplementary Table 5: Latent Class Summary with Item Response Probabilities.**

| Latent Class | D1 | D2 | D3 | D4 | D5 | D6 | D7 |
|---|---|---|---|---|---|---|---|
| **High adherence** | 0.524 | 0.818 | 0.928 | 0.75 | 0.877 | 0.447 | 0.922 |
| **Low adherence** | 0.136 | 0.000 | 0.394 | 0.143 | 0.433 | 0.080 | 0.443 |

**Supplementary Table 6: Showing the percentage of adherence and non-adherence to prescribed diet with demographic variables and BMI.**

| Variables | Low Adherence | High Adherence | Chi Sq. p-value |
|---|---|---|---|
| **Overall** | 33 (31.7) | 71 (68.3) | --- |
| **Sex** | | | |
| Male | 25 (35.7) | 45 (64.3) | 0.304 |
| Female | 8 (23.5) | 26 (76.5) | |
| **Alcoholic** | | | |
| No | 24 (30.0) | 56 (70.0) | 0.658 |
| Yes | 9 (37.5) | 15 (62.5) | |
| **Smoking** | | | |
| No | 32 (31.7) | 69 (68.3) | 0.999 |
| Yes | 1 (33.3) | 2 (66.7) | |
| **BMI** | | | |
| Normal | 12 (30.0) | 28 (70.0) | 0.861 |
| Underweight | 0 (0.0) | 2 (100.0) | |
| Overweight | 2 (22.2) | 7 (77.8) | |
| Obese I | 15 (34.9) | 28 (65.1) | |
| Obese II | 4 (40.0) | 6 (60.0) | |

**Bonferroni corrected p<0.0125 for significance; values n (%).*

**Supplementary Figure 1. Biplots showing quality of representation (cos²) of variable categories across all MCA dimension pairs**

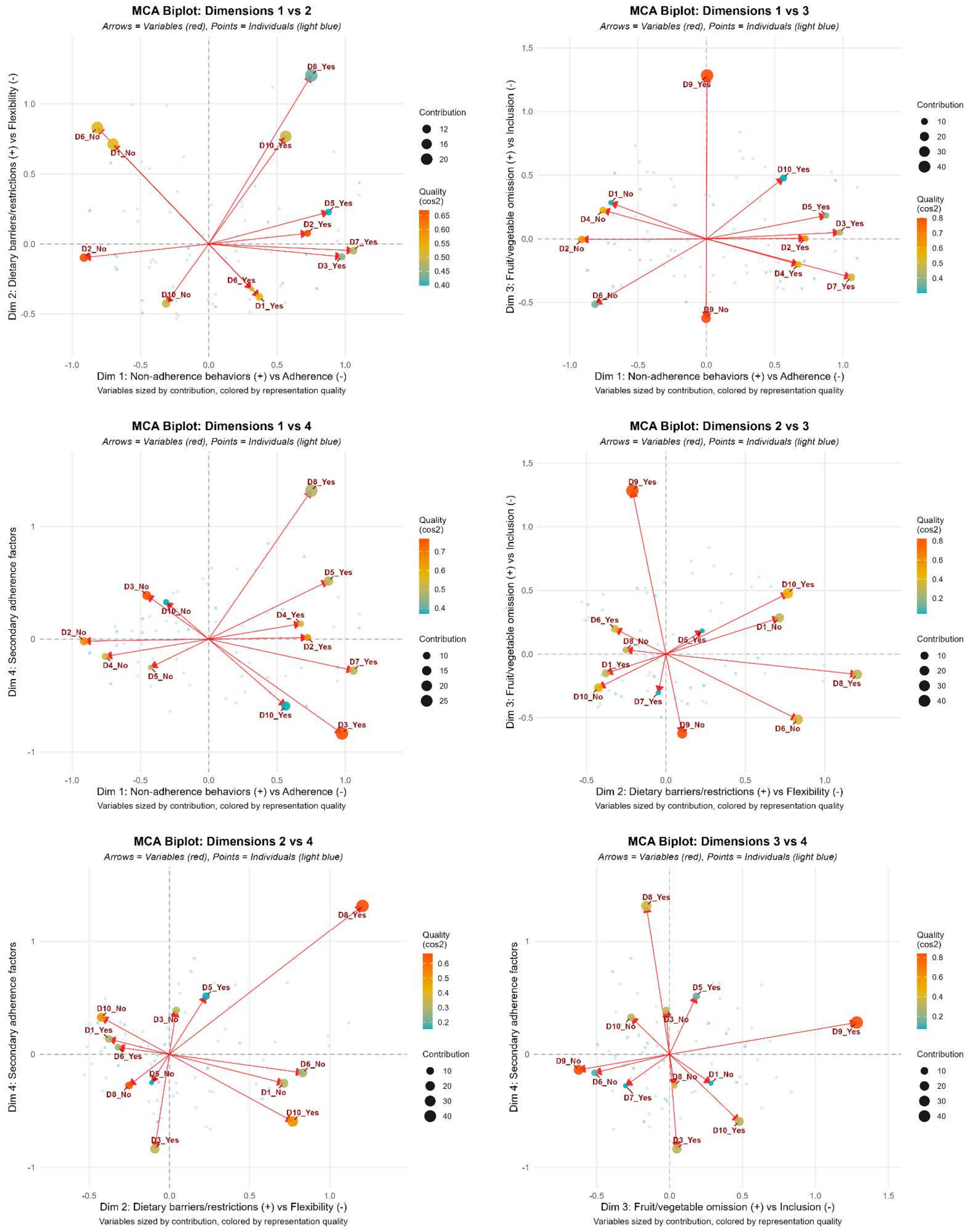


*Here "no" corresponds to adherent and "yes" to non-adherent for that question.*

**Supplementary Figure 2. Plot shows association maps of variables from MCA (all dimension pairs).**

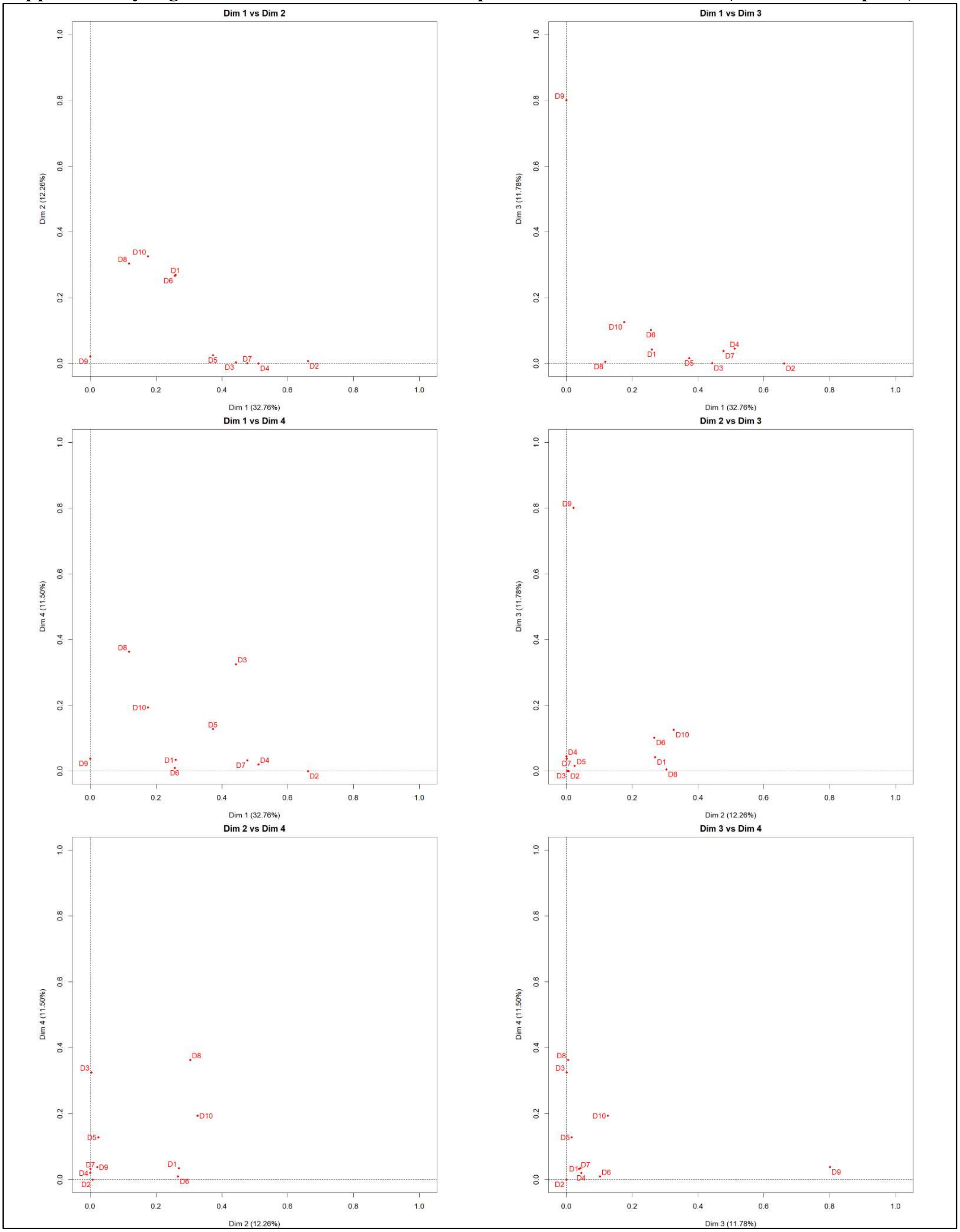

**Supplementary Figure 3. Heatmap of standardized residual covariances from the confirmatory factor analysis (CFA) model.**

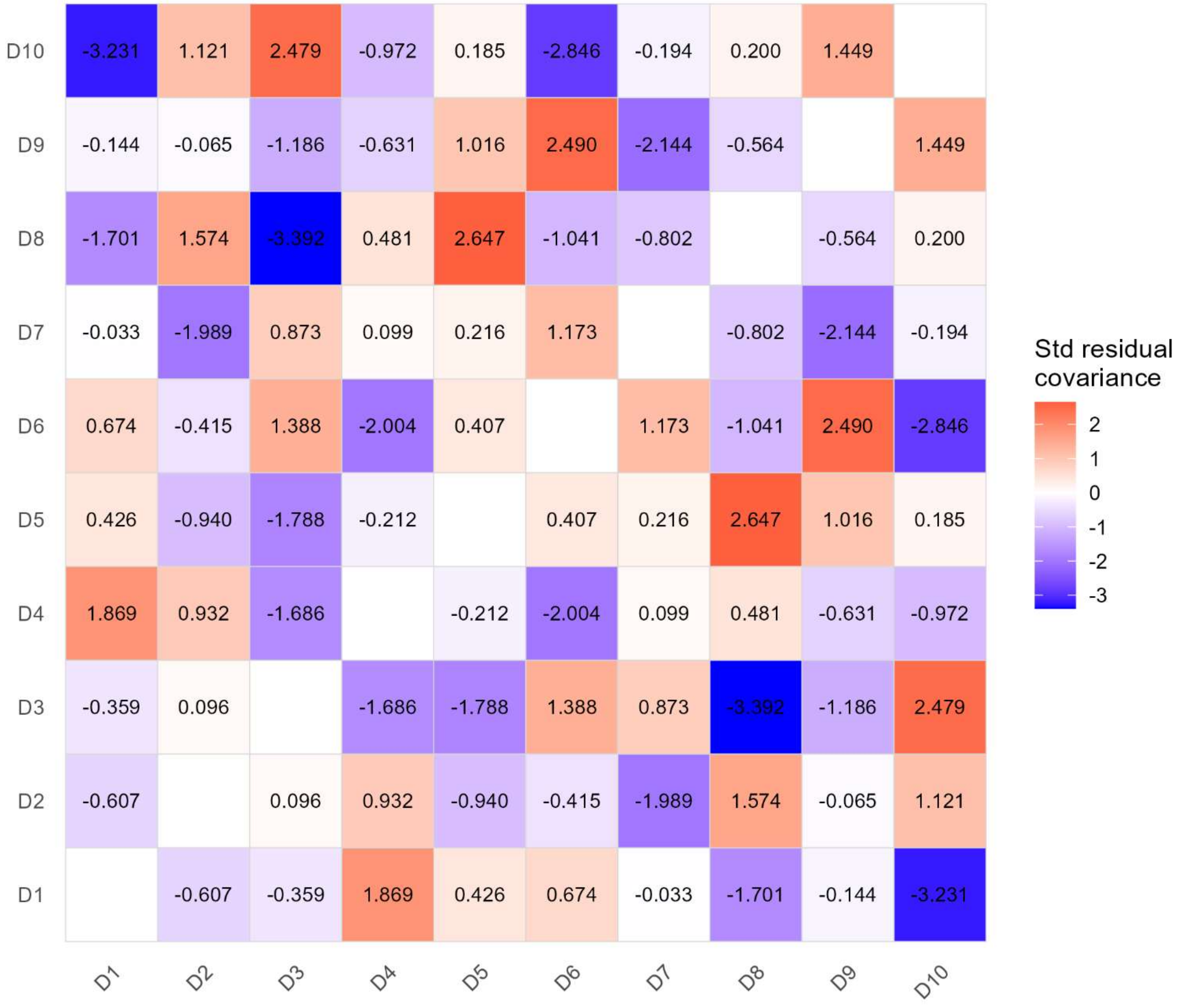

**Supplementary Figure 4. Plot shows item response theory parameter comparison for 7-Items and 10-Items Model.**

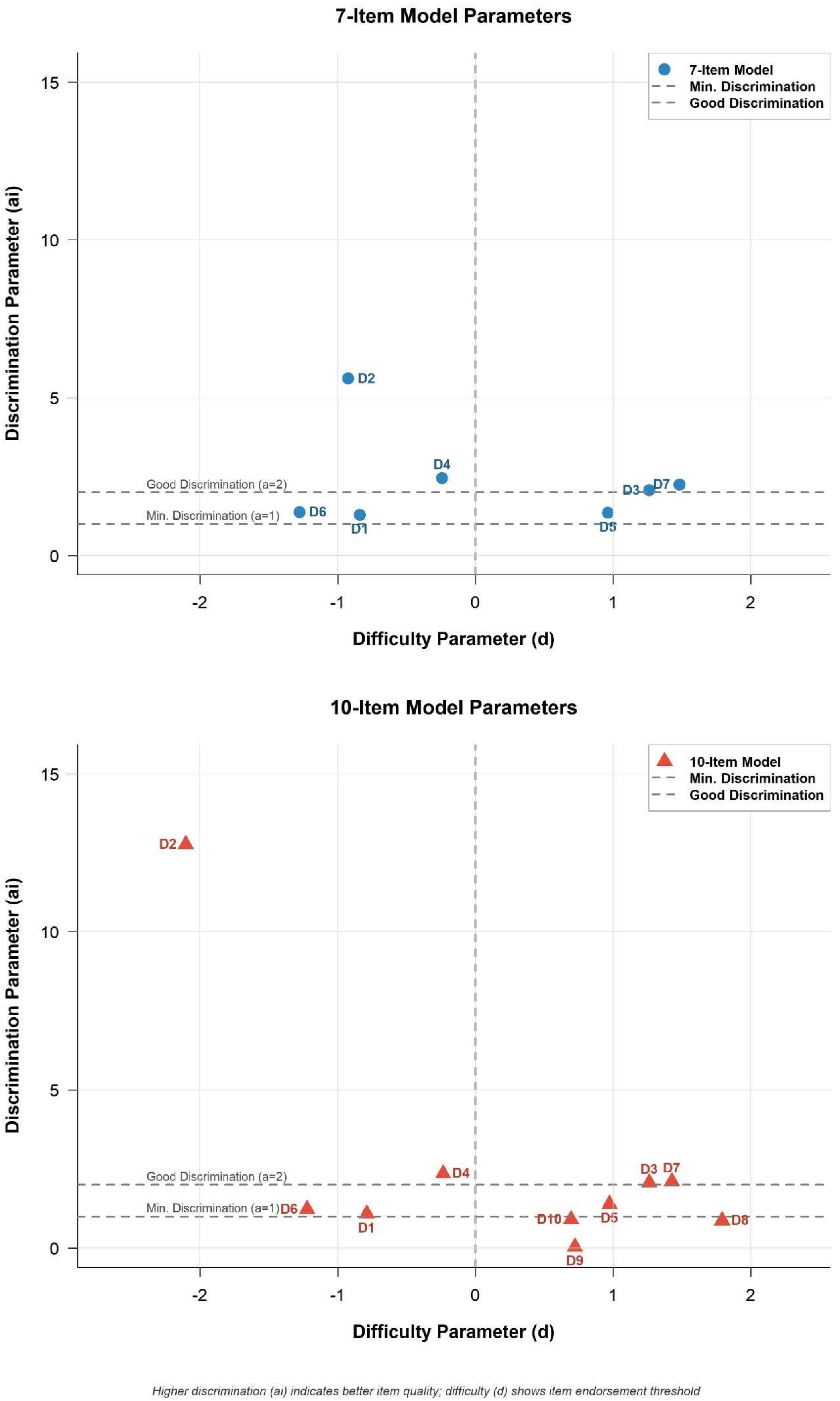
